# Gods Lockdown: the effect of the April 2022 Colorado low blizzard on SARS-Cov-2 transmission in the midwest United States

**DOI:** 10.1101/2023.11.01.23297908

**Authors:** Stephenson Strobel, Michael Daly

## Abstract

**Background:** Lockdowns have been used as a primary non-pharmaceutical intervention to stop transmission of COVID19. There are many issues with interpreting the causal effects of most of these intentional, policy driven interventions. We leverage a natural experiment to avoid many of these issues to better understand the direct effects of lockdown like conditions on COVID19 transmission.

**Methods:** We exploit a blizzard that interrupted activity across several midwestern states in April 2022. This blizzard broke records for snowfall and caused economic disruption. We leverage this to create control and treatment counties that were more or less affected by the snowfall. We demonstrate effects using event studies comparing these treatment and control counties.

**Results:** We find that mobility within treatment counties was severely curtailed as a result of the blizzard relative to control counties. We find cumulative declines in the number of COVID19 cases per country by 400 and cumulative declines in COVID19 deaths by 1 per county over the 30 days after the storm. We find declines in by one per hospitalization due to COVID19.

**Conclusions:** The April 2022 blizzard caused disruption in activity across the midwest United States akin to a lockdown. It reduced the number of COVID19 cases, deaths, and hospitalizations in treatment counties relative to control counties suggesting that similar policies do limit transmissions of SARS-Cov-2.

## Background

Lockdowns are one of the primary non-pharmaceutical interventions (NPI) used during the COVID19 pandemic to prevent spread of SARS-Cov-2. They were meant to prevent contact between infected and uninfected individuals and break an escalating chain of transmission that overwhelmed hospital systems, and caused major morbidity, and mortality. Such lockdowns typically curtailed economic activity and social mobility by instituting curfews and closing non-essential businesses ^1,2^. Several countries and subnational jurisdictions including those in the United States, China, Italy, and Singapore implemented lockdowns with varying degrees of harshness^1,3–6^.

There have been previous studies that have evaluated the effectiveness of COVID19 lockdowns. However, there are two major issues with a causal interpretation of these interventions; first, underlying characteristics of the population may be driving the observed effects^3,4,7^. Places with lockdowns may be more likely to have populations that are more cautious in nature and more likely to comply with the restrictions. It is not only the lockdown but the unobserved characteristics of this population that are causing declines in cases. Second, lockdowns for COVID19 were not done in isolation of other policies. In most cases, there were a host of other interventions implemented to curb transmission such as masking, medical system limitations and promotion of social distancing^5^. It is unclear whether these analyses are measuring the true effect of the lockdowns, or a contaminated effect caused by unobserved population heterogeneity or alternate NPIs.

To understand the causal effect of a lockdown on COVID19 transmission in a community, it would be ideal to randomize lockdowns to jurisdictions. Ethical concerns and logistics prohibit this and so we exploit a natural experiment that occurred in the US that approximates randomizing lockdowns. In April 2022, a Colorado Low weather pattern developed over the Rocky mountains and moved north into Canada producing up to 45 inches of snow in North Dakota, South Dakota, Montana, and Minnesota^8^. This necessitated the closing of schools and severely curtailed travel to work, transit use, and public activity. Moreover, this first blizzard occurred over several days and a subsequent blizzard occurred in these states just one week later^9^.

We examine this April 2022 blizzard as an approximation of a lockdown on certain treatment counties but not control counties that were less affected by the snowfall from the storm. The benefit of this is twofold. First, we can examine a roughly homogenous group of counties in the United States. Second, the event of the blizzard is unlikely to induce other NPI related behaviours like changes in masking. We assess its impact on economic activity, COVID19 cases and hospitalizations using a differences-in-differences econometric strategy. We show that the blizzard did cause reductions in mobility and economic activity as evidenced by Google data. Cases and deaths both declined in treatment counties relative to control counties just after the blizzard. Hospitalizations also declined. This provides evidence on the benefits of lockdowns on infectious disease transmission for policy makers interested in their use during the COVID19 pandemic and in future pandemics.

## Methods

### Context

We exploit a weather complex as a natural experiment that forced certain counties in the midwestern United States to experience lockdown-like conditions due to the amount of snow brought on by a blizzard. This Colorado low-pressure weather system moved across the midwestern United States during April 2022 causing tornadoes and other severe weather phenomena in the south and heavy snowfall conditions in the north. We focus on these latter weather patterns which were so severe that several weather records were broken across Montana, South Dakota, North Dakota, and Minnesota^10^.

Billings, Montana recorded the snowiest April day since 1955 and Bismarck, North Dakota recorded their snowiest April month ever on record^8,11^. Multiple counties across North Dakota broke snowfall records and much of the interstate highway system in North Dakota was shut for safety reasons^12^. At least one person was killed while traveling by car during this period^13^. This first wave of snowfall on April 12-14 was then proceeded by a second April blizzard from April 22-24 which also significantly impacted travel especially in North Dakota^9^.

### Data

We combine data with our main variables of interest at the county level from four different data sources. First, we create treatment and control counties using data on the snowfall conditions from April 12 to April 24, 2022 over the period when these snowstorms were occurring. These data are at the county level and are sourced from the National Centers for Environmental Information daily US snowfall and snow depth data sets^14^. These include information on the snowfall at all weather stations within the states of Montana, South Dakota, North Dakota, and Minnesota which we restrict our analysis to. Given some sparsity of weather stations in the midwest United States we make inferences about the treatment status of a county that has no data by the snowfall in the surrounding counties. We arbitrarily designate a county as a treatment county if they cumulatively receive over 10 inches of snowfall during the period of the two blizzards. As a descriptive exercise we provide some evidence on the population within treatment and control counties using data from the 2020 US census.

Second, we use Google community mobility data to demonstrate a first stage effect on social mobility within these treatment counties relative to control counties^15^. This outcome reflects mobility in a county as a percent change relative to April 2020. Thus outcomes can be interpreted as a decline or increase in mobility as a percent of this baseline date. Google is able to estimate whether activity is occurring within certain designated locations such as residential or workplace areas. We focus on mobility in these two area types as our key first stage outcome that provides evidence that behaviour changed similar to other lock-down events.

Third, we use county level COVID19 case and death data collected by the New York Times^16^. These data are the daily case count and death data that have been pieced together from public health reports, journalist interactions with officials, and news conferences. They consist of a daily cumulative count of the number of confirmed and probable cases and deaths due to COVID19 in a county. We provide evidence using both the cumulative cases and deaths and new cases and deaths which we create as outcomes. There are occasions in this data where cumulative cases decline likely due to recording issues; where this occurs we label these dates where “new” cases or deaths are negative as missing data.

Finally, we examine effects on hospital related outcomes. We do this to assess COVID19 specific outcomes like the number of adult beds and adult ICU beds used by these patients. We also examine the overall admission rate as a check on whether results are driven by health effects of the storm that were not related to COVID19.

These data are sourced from HealthData.gov which collects supply and demand for beds at all hospitals registered with the Centers for Medicare & Medicaid Services^17^. These data are at the facility level and are observed each week.

We are interested in four outcomes. First, we are interested in the amount of snowfall in a county during the blizzard period; this dictates how we create our treatment and control group of counties. Second, we are interested in the mobility of individuals within those treatment and control counties as a first stage on whether the storm created lockdown like conditions. Third, we are interested in the new and cumulative number of total cases and deaths due to COVID19 within a county by their treatment status.

Finally, we are interested in the impacts on hospital bed use by COVID19 and non-COVID19 patients for hospitals within treatment and control counties.

### Identification strategy

To identify causal impacts we employ Callaway-Sant’ana difference-in-differences estimators to evaluate pre-post storm effects among treatment and control group counties^18^. Given that the effects of reducing COVID19 transmission may occur later than an event that interrupts transmission we specifically look at event analyses. In the case of the mobility and case or death data these are at the county level and are observed daily. We estimate the following

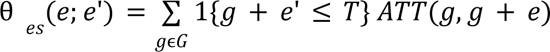

where θ is the estimate of the average effect of being in a treatment county *e* days after the snow storm. G denotes the time of first treatment which is the April 12 date and is the same for all treated counties. This is our window of observation for effects to develop and for us to evaluate pretrends. We estimate the following average treatment effect:

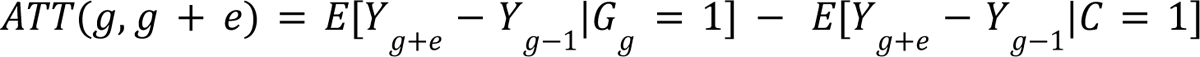

where we compare the treated group of counties (G=1) to the control group of counties (C=1). Outcomes, *Y*, are as specified previously. In these regressions we cluster standard errors at the county level. We do not control for any other variables and so these are regression adjusted estimates^19^.

For regressions at the hospital level our unit of time is at the week level and our observations are at the hospital. In these cases we observe for six weeks pre and post the April 12 week of initial treatment. We cluster our standard errors in these regressions at the county level in these regressions as we suspect that treatment occurs at this level as well^20^.

For causality to hold in event analyses, there must be no treatment anticipation between control and treatment counties and there must be no other treatment that would contaminate effects of the storm. The former can be tested in these events by examining the pretrends of treatment and control counties. We demonstrate these in our results by plotting all events^21^.

## Results

We first document snowfall from the April 2022 blizzard and how it affects counties in figure 1. The initial blizzard tracked through the center of the four states in our sample from April 11 to April 14 and a subsequent blizzard moved through the same area from April 22-24, 2022. At its epicenter along the Montana-North Dakota border, these two blizzards produced a cumulative 30 to 50 inches of snow. Figure 1 also demonstrates how this creates our treatment and control counties along this track of the blizzard.

**Figure 1:**
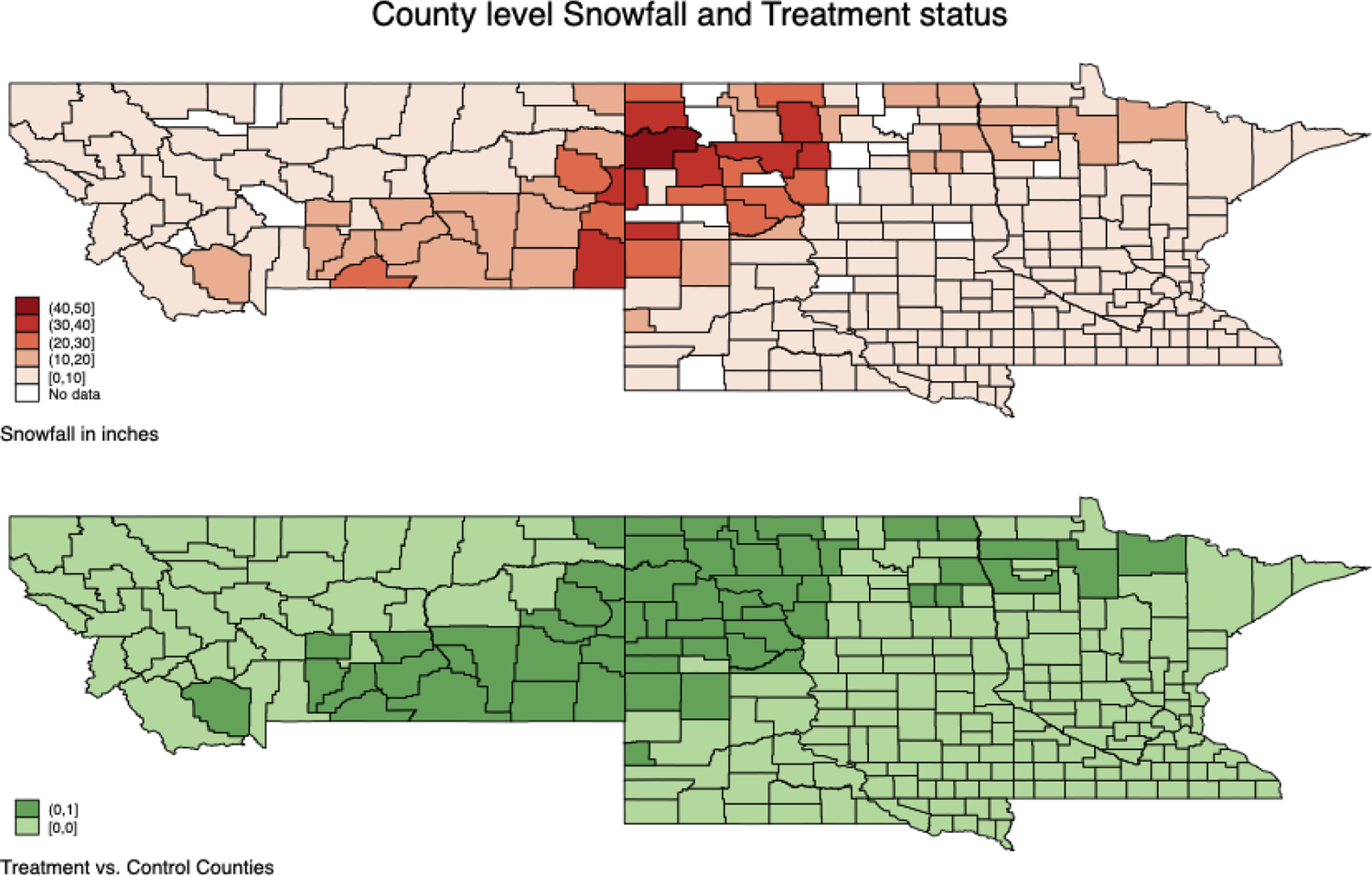
Maps of the counties within Montana, North Dakota, South Dakota and Minnesota overlaid by snowfall amount during the period of April 11 to April 24, 2022. Cumulative snowfall over 10 inches designates a county in the treatment group relative to control counties that had less snowfall.

Table 1 documents the descriptive characteristics of treatment and control counties by state. These show that treatment counties tend to have smaller populations and have more of them living in rural areas. We find low case counts in treatment and control counties prior to the storm in April. The exception is in Minnesota where daily new cases are relatively high in both treatment and control counties. Our treatment counties have less of a proportion of their population within urban areas and have smaller populations which point to their being more rural than control counties.

**Table 1:**
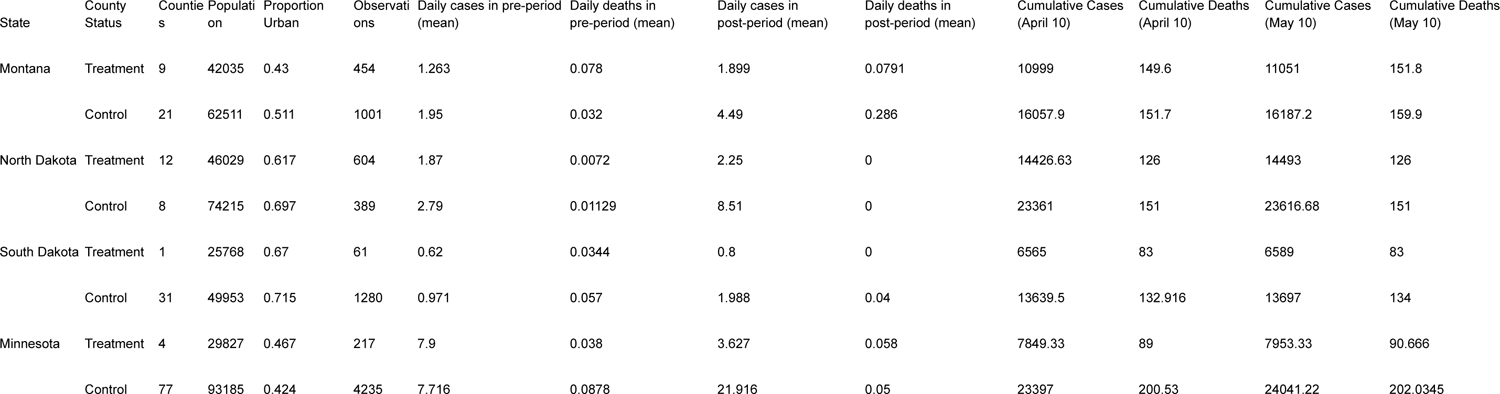
Descriptive statistics of the treatment and control counties by state and pre and post period.

We next assess first stage effects on mobility using Google data. Figure 2 demonstrates changes in mobility scores around the period of the initial storm for workplaces and residences relative to April 2020. These changes are relative and should be interpreted as a difference in our treatment counties relative to control counties. We find large declines in mobility at workplaces and corresponding large increases in mobility at residents around the time of the initial storm. These last approximately 4-5 days. We find similar signed impacts of the second wave of the blizzard one week later, although the effects are of lower magnitude. Our pre-trends in this data demonstrate little sign of differential anticipatory behaviour between control and treatment counties. We also find strong correlations between differences in mobility score and the amount of snowfall by date suggesting our treatment and control groups capture an element of treatment by this blizzard (figure e1).

**Figure 2:**
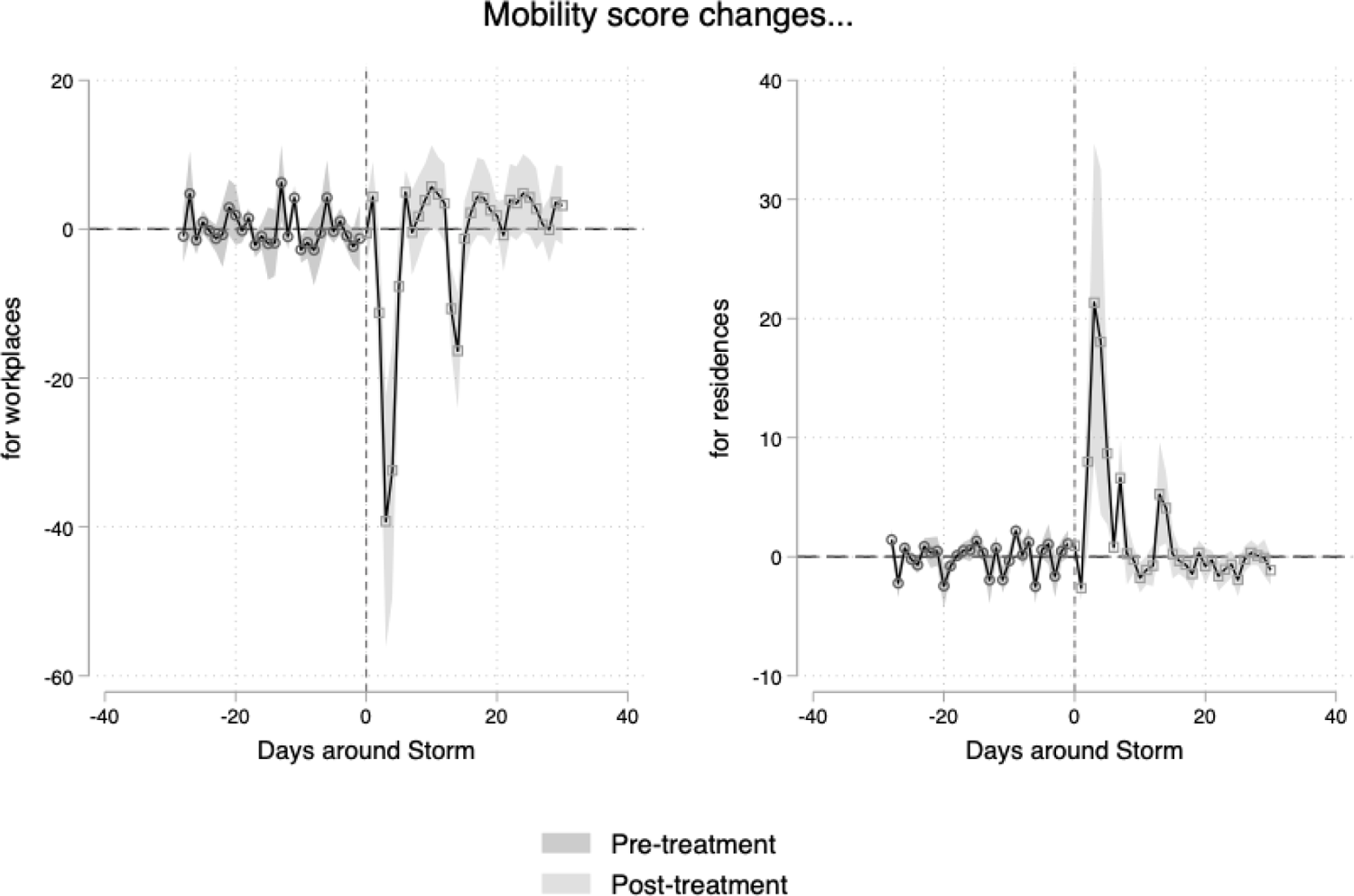
Event analyses showing mobility scores in treatment counties relative to control counties for workplaces and residences. Day 0 is April 11, 2022 or when blizzard conditions began. Mobility scores are relative to a period in April 2020.

We can compare the impacts of these storms to other lockdown episodes that are in the Google mobility data. For example, in figure e2, we demonstrate the time series of the same daily mobility data for a period around March 2020 when several US states instituted the first stay-at-home orders and closed non-essential businesses. We find that the impact of the initial storm is similar in magnitude to these lockdowns although the persistence is much more limited as would be expected with a passing storm.

We turn to examining county level COVID19 case and death data. Figure 3 demonstrates the impacts of the storm on new and cumulative COVID19 cases and deaths. Over the 30 days after the storm, we find a reduction in the daily new cases by approximately 25 to 50 per county and a cumulative decline in cases by 200-400. These effects develop immediately after the initial storm and continue for our 30 day window of observation.

**Figure 3:**
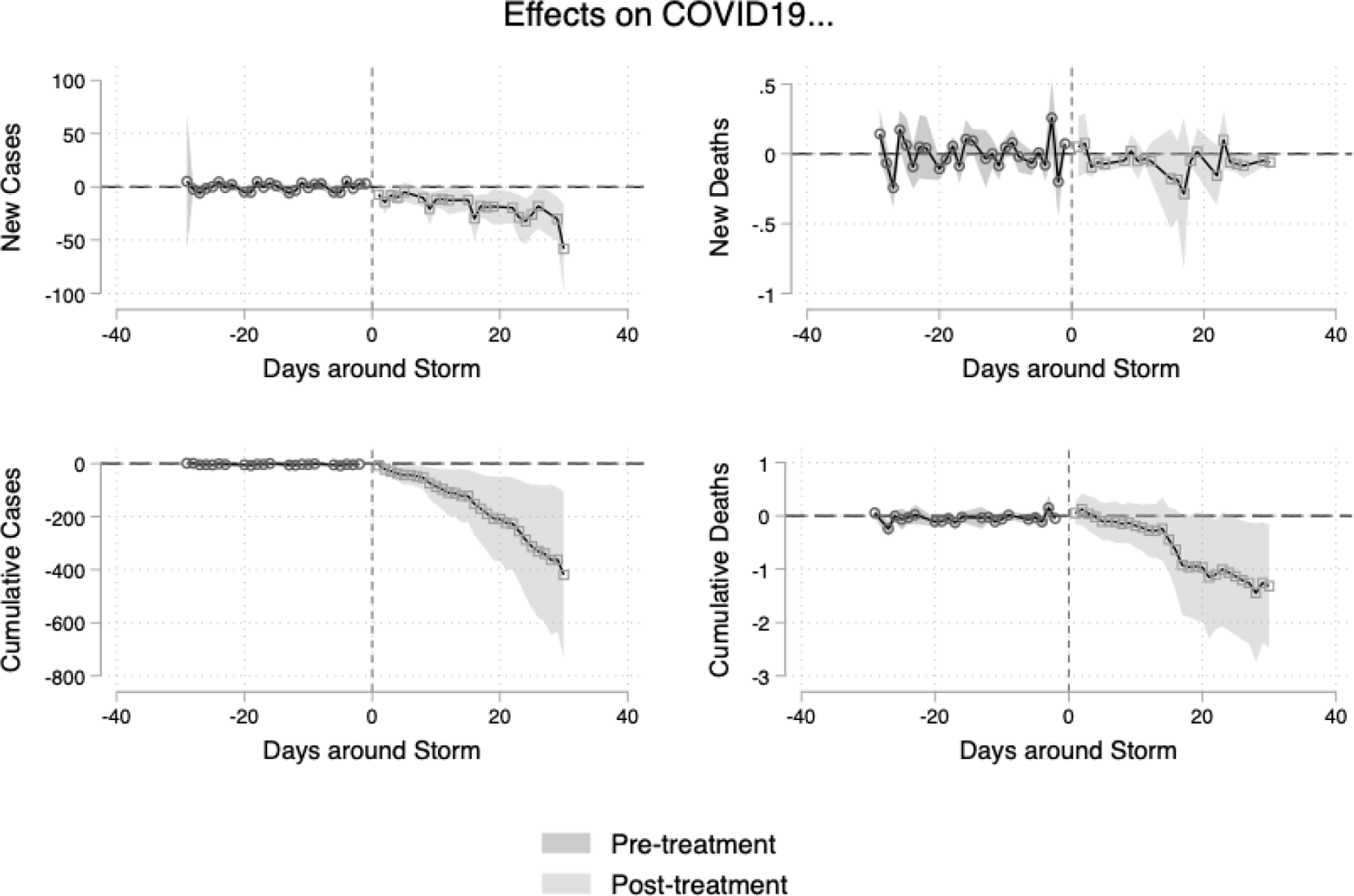
Event analyses showing new and cumulative cases and deaths due to COVID19. Day 0 is April 11, 2022 or when blizzard conditions began.

We find similar declines in the number of new deaths and cumulative deaths in treatment counties relative to control counties. At its peak, this results in a reduction in the number of new daily deaths due to COVID19 by 0.25 approximately two weeks after the initial blizzard. This results in a cumulative decline in the number COVID19 deaths by one case per county with the majority of this effect plateauing two weeks after the initial storm. Our pre-trends in these data also demonstrate little sign of differential anticipatory behaviour between control and treatment counties.

Finally, we assess impacts of the storm on hospital bed use. Figure e3 demonstrates impacts on the number of hospital beds used around the time of the storm for both COVID19 and all patients. We find modest declines in the number of beds used by COVID19 patients in the weeks after the storm occurred in treatment counties relative to control counties. There are no impacts on COVID19 ICU bed use or the number of overall patients in all beds or ICU beds. We find some suggestion that there may be pretrends in COVID19 hospitalizations that are present five to six weeks before the storm but dissipate by the immediate pre-period before the blizzard.

## Discussion

We find that the April 2022 Colorado low blizzard had impacts on areas in the midwest United States with particularly high levels of snowfall. Relative to counties with less snowfall, these treatment counties saw decreases in Google mobility scores that were similar in magnitude, although shorter-lived, to previous lockdown style policies. In conjunction with these declines in mobility there were declines in the number of reported cases and number of deaths in these treatment counties. There were also declines in the number of hospitalizations due to COVID19 in hospitals in these counties.

These results demonstrate that disrupting social activity like that which occurs during a lockdown can reduce case transmissions and prevent deaths. However they must be interpreted with some caveats; first, the type of lockdown that this event approximates is a short, sharp decline in activity. These results can best be applied to “circuit-breaker” style lockdowns that were being suggested by public health officials at the beginning of 2022 to stop out of control COVID19 transmission^5,22^. Our example is even shorter in duration than the types of lockdowns implemented in places like Singapore and the UK. Further, our results poorly approximate the type of lockdowns that curtailed activity for much longer periods of time at the onset of the pandemic.

We also caution about how these effects develop over time in our data. We find an immediate significant decline in COVID19 cases after the first blizzard hit our treatment counties. Given that biological studies of SARS-Cov-2 suggest that symptoms develop after several days of being infected we suspect this initial drop is due to fewer cases presenting for medical evaluation and tabulation^23^. This initial decline is unlikely to be a true change in the number of COVID19 cases. However this decline continues over several weeks in new cases and cumulative cases suggesting true effects on COVID19 transmission even as economic activity normalized after the storm.

These results also need to be contextualized in terms of their magnitude. We find statistically significant and clinically significant impacts on cases. Our results are consistent in sign to most of the current literature on lockdowns^1,3^. We find reductions in the range of 25 fewer new cases per treatment county per day and cumulative reductions by 400 cases by 30 days after the blizzard. How do we assess these impacts with regards to our descriptive statistics? We suspect that rather than stop an ongoing outbreak of COVID19, the storm prevented an outbreak in most treatment counties.

Both treatment and control counties display minimal new cases in the weeks prior to the storm. After the storm though, control counties, especially those in North Dakota, that did not experience impacts of the blizzard saw large increases in COVID19 case counts. The exception to this was in Minnesota where treatment counties were experiencing relatively high new daily case counts which then collapsed after the blizzard.

However, while statistically significant, we find more clinically modest reductions in COVID19 deaths. We find that in treatment counties, the effects are to reduce the cumulative number of deaths by one relative to treatment counties. Overall, this would suggest that the blizzard prevented 22 COVID19 deaths during the 30 days after its impact. We cannot make any inferences about the impact of this blizzard in particular, or the impact of circuit breaker lockdowns on lost economic activity in general. However, if our estimates reflect the benefits of a lockdown and the value of a statistical life is in the range of 7 to 10 million USD^24^, a hypothetical lockdown in these counties would have to interrupt approximately 154 to 220 million USD in economic activity for costs to outweigh the benefits of lives saved. This back of the envelope calculation does not include prevention of morbidity in persons who survive and does not take into account the age-structure of COVID19 deaths which predominantly affects older individuals^25,26^.

The major limitations of these results relate to assumptions built into the event analyses. We find that our parallel trends assumption is relatively strong as evidenced by pretrends in our event plots. This suggests little differential anticipation of treatment by the blizzard. However, mobility is unlikely to be the only outcome affected when a blizzard hits a county. We find little evidence that hospitalizations for non-COVID19 patients changes in conjunction with the blizzard suggesting against treatment effects that negatively affect health overall. We also suspect that our results are heavily contextual and would be different in a pre-vaccine environment. Their applicability also depends on the virulence of the disease and the policy environment which are almost certain to be different during other pandemics. We would urge caution with applicability in these alternative scenarios.

## Conclusion

We examine the impacts of the April 2022 blizzard on Sars-Cov-2 transmission in the midwestern United States. We find evidence that the short and sharp decline in mobility caused by the storm had impacts on cases, deaths, and hospitalizations due to COVID19. Further research should characterize the trade-offs that necessarily occur by curtailing economic activity given our estimates of the prevented cases and deaths as a result of this blizzard created lockdown.

## Data Availability

All data produced in the present study are available upon reasonable request to the authors and will be provided to the relevant journal upon publication.

---

**Figure e1:**
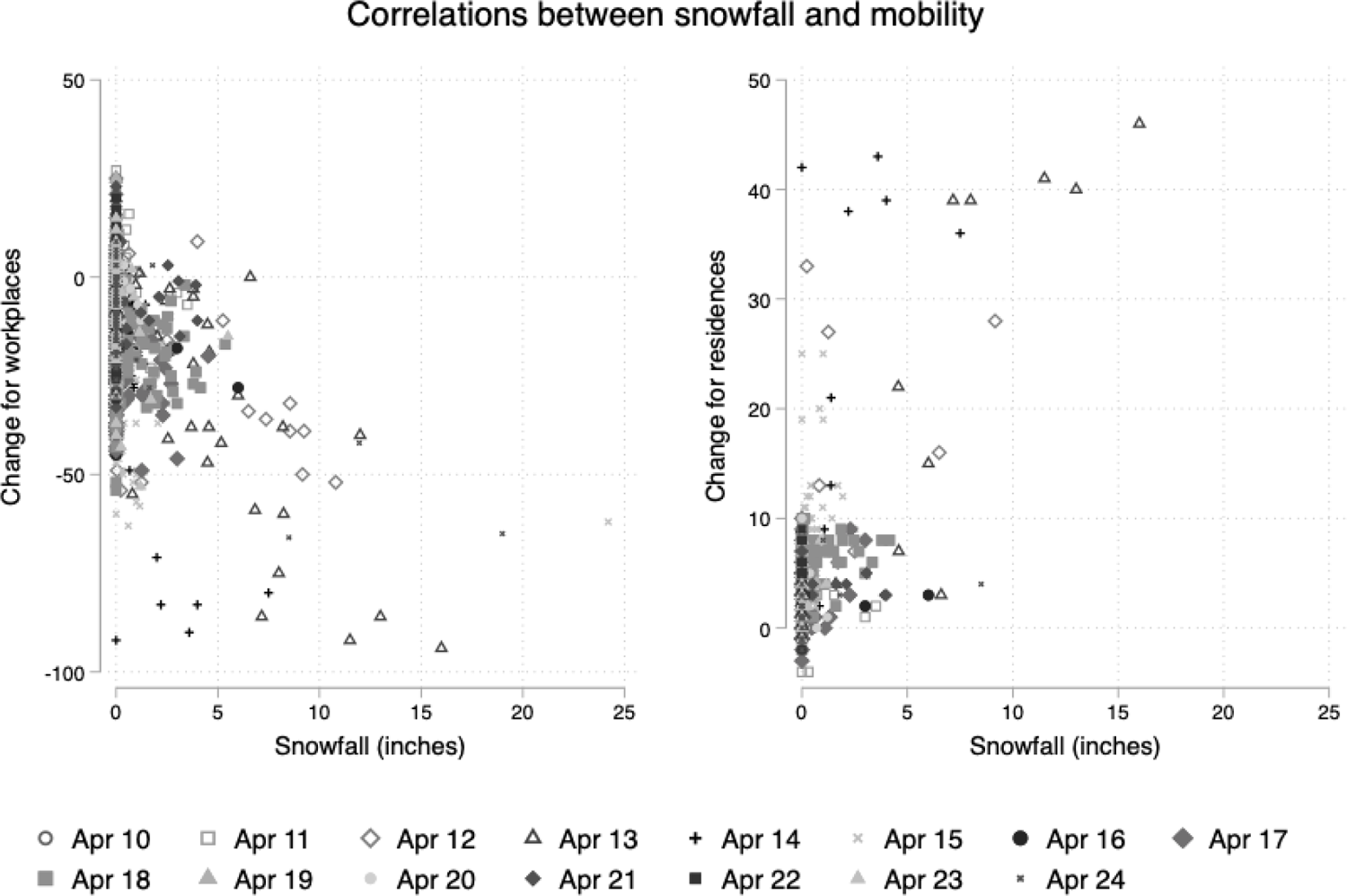
Correlations between the daily snowfall and mobility within a county by date for residential and workplace scores. Note that the blizzard occurred on April 12-14 and 22-24.

**Figure e2:**
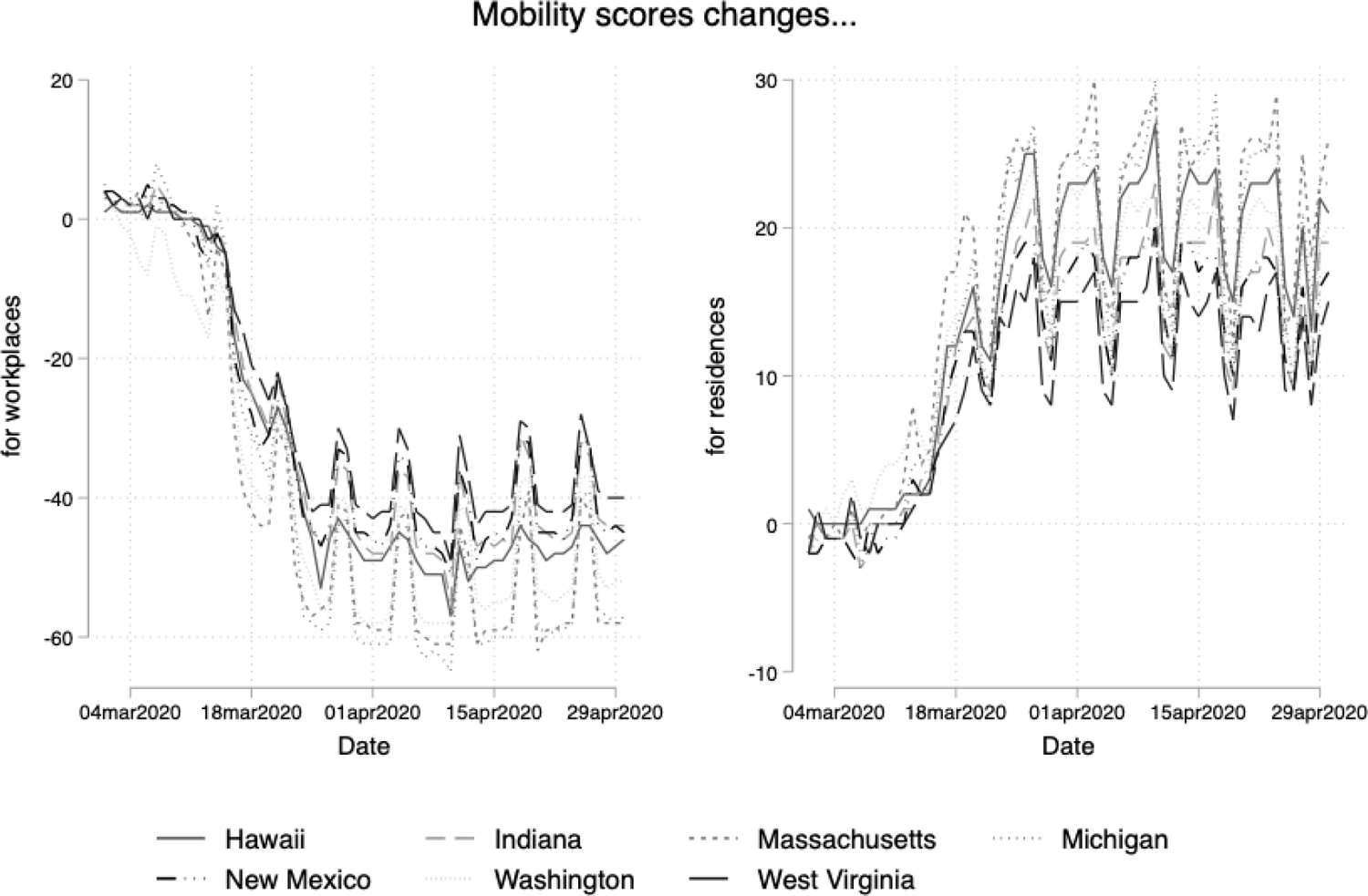
Mobility score changes for several select states that instituted lockdown like policies during the period of March 2020.

**Figure e3:**
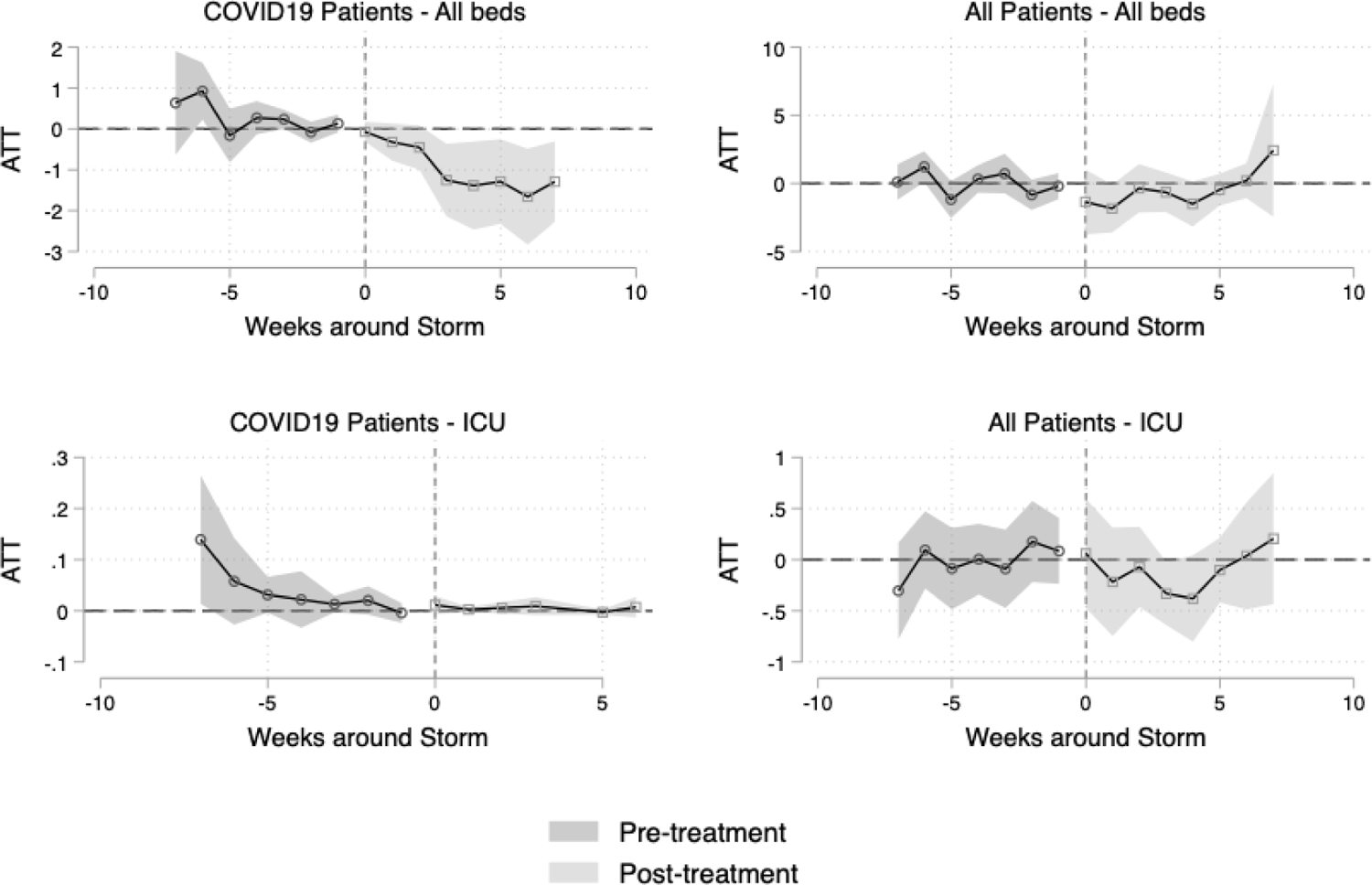
Event analyses showing the number of beds in use for COVID19 patients and all patients for hospitals in treatment counties relative to control counties. Week 0 coincides with April 12, 2022 or approximately when blizzard conditions began.

